# Diagnostic power of resting-state fMRI for detection of network connectivity in Alzheimer’s disease and Mild Cognitive Impairment: A systematic review

**DOI:** 10.1101/2020.08.28.20182931

**Authors:** Buhari Ibrahim, Nisha Syed Nasser, Normala Ibrahim, Mazlyfarina Mohamed, Hasyma Abu Hassan, M. Iqbal Saripan, Subapriya Suppiah

## Abstract

Resting state fMRI (rs-fMRI) detects functional connectivity (FC) abnormalities that occur in the brains of patients with Alzheimer’s disease (AD) and Mild Cognitive Impairment (MCI). FC of the default mode network (DMN), which is involved in memory consolidation, is commonly impaired in AD and MCI. We aimed to determine the diagnostic power of rs-fMRI to identify FC abnormalities in the DMN, which help to distinguish patients with AD or MCI from healthy controls (HCs). We searched articles in PubMed and Scopus databases using the search terms such as AD, MCI, resting-state fMRI, sensitivity and specificity through to 27th March 2020 and removed duplicate papers. We screened 390 published articles, and shortlisted 12 articles for the final analysis. The range of sensitivity of DMN FC at the posterior cingulate cortex (PCC) for diagnosing AD was between 65.7% - 100% and specificity ranged from 66 - 95%. Reduced DMN FC between the PCC and anterior cingulate cortex (ACC) in the frontal lobes was observed in MCI patients. AD patients had impaired FC in most regions of the DMN; particularly the PCC in early AD. This indicates that DMN’s rs-fMRI FC can offer moderate to high diagnostic power to distinguish AD and MCI patients. fMRI detected abnormal DMN FC, particularly in the PCC that helps to differentiate AD and MCI patients from healthy controls (HCs). Combining multivariate method of analysis with other MRI parameters such as structural changes improve the diagnostic power of rs-fMRI in distinguishing patients with AD or MCI from HCs.

## 1 Introduction

Alzheimer’s disease (AD) is a neurodegenerative disorder that is characterized by a progressive decrease in cognitive function compared to baseline performance level in one or more cognitive domains that can interfere with the ability to independently carry out activities of daily living (American Psychiatric Association, 2013). Resting state functional magnetic resonance imaging (rs-fMRI) is a neuroimaging tool used to study the aberrations in the functional activity of different brain networks, which normally occurs in AD and its prodromal condition, mild cognitive impairment (MCI) (Chen et al., 2011). The functional connectivity (FC) of brain networks refers to interregional synchrony, as detected from low frequency fluctuations in the blood oxygen level dependent (BOLD) fMRI sequence (Lee, Harrison, & Mechelli, 2003). FC is studied using different molecular imaging techniques such as electroencephalography (EEG), positron emission tomography computed tomography (PET/CT) and fMRI. Of these techniques, fMRI remains as the most widely used because of the relative simplicity of usage, safety, non-invasive nature and high spatial resolution provided by the technique (Mier & Mier, 2015).

Default mode network (DMN) is the commonest brain network studied by rs-fMRI and is involved in memory consolidation tasks. It comprises of the precuneus (Prec), posterior cingulate cortex (PCC), retro-splenial cortex, medial, lateral and inferior parietal cortex (MPC), (LPC), and (IPC), medial prefrontal cortex (mPFC) and the medial temporal gyrus (MTG) (Mohan et al., 2016; Wermke, Sorg, Wohlschläger, & Drzezga, 2008). AD patients suffer from impaired DMN connectivity (Grieder, Wang, Dierks, Wahlund, & Jann, 2018). There has been consistent evidence of decreased FC in the DMN of AD patients in comparison with HCs, especially between the posterior part of the cerebral cortex (Prec and PCC) and anterior parts, for example the anterior cingulate cortex (ACC) and mPFC (Brier et al., 2012; Gili et al., 2011; L Griffanti et al., 2015). The observed decline in FC in areas within the DMN have also being reported among MCI patients (Cha et al., 2013; Filippini et al., 2009; Ouchi & Kikuchi, 2012). This indicates that fMRI detected changes in the DMN can be a non-invasive diagnostic tool for diagnosing AD.

There are several methods to analyse rs-fMRI data, namely the seed-based analysis (SBA), the independent component analysis (ICA) and the graph theory analysis (GTA). SBA or small region of interest (ROI) analysis enables temporal correlations to be made between seed regions. The SBA investigates the FC of a given brain region by correlating the brain regions’ resting-state time series with the time series of all other regions resulting in the creation of a FC map identifying the FC of the pre-defined brain regions (Jiang, He, Zang, & Weng, 2004). The simplicity and straight forwardness of this seed-dependent analysis coupled with the clarity of the FC map, makes it popular among the users of the rs-fMRI modality (Buckner et al., 2009). Nevertheless, the knowledge from a FC map is restricted to the FC of a pre-defined region, making it hard to analyse correlations of FC in whole brain regions.

In contrast to SBA, ICA is free from any pre-defined seed region selection, which means one does not have to pick a seed or reference area beforehand. Hence, the entire BOLD signal is broken down to produce separate time courses and related spatial maps (De Luca, Smith, De Stefano, Federico, & Matthews, 2005). The resultant components are independent of one another. Due to its ability to accommodate whole-brain FC analysis, ICA is favoured over SBA. Nevertheless, the disadvantage of ICA is that often there is difficulty in differentiating useful signal from noise and variations in the separate components. Thereby, this causes challenges in making between group comparisons using ICA (Fox & Raichle, 2007). Interestingly, both SBA and ICA can produce similar results if they are run in different experimental set-ups.

Alternatively, GTA looks at the overall brain network structure with specific spatial information. Here, the BOLD signal undergoes spatial parcellation using a topological mapping of the entire brain and the relationships between all pairs of activated regions are determined. This is achievable by forming a 2×2 matrix of nodes versus edges of the activated regions. A ‘node’ is a defined area in the brain, whereas ‘edge’ signifies the direct and indirect links between two defined nodes. In this way, the brain is considered as a single complex network where several global and local network topologies such as path length, modularity, and efficiency can be measured (Rubinov & Sporns, 2010). Consequently, resting state networks identified using SBA and ICA can be confirmed with GTA.

Furthermore, studies have analysed the relationship between the FC of the DMN, Mini-Mental State Examination (MMSE) test scores, and the development of disease among amnestic MCI (aMCI) and AD patients and compared them with HC subjects (Cha et al., 2013; Liao et al., 2018; Liu et al., 2016). aMCI and AD groups had decreased FC in left PCC and left parahippocampal gyrus as compared to HC subjects (Liu et al., 2016; D. Zheng et al., 2018). Only AD patients were identified with increased FC at the right middle frontal gyrus (MFG), which was interpreted as a compensatory neural mechanism in response to the impairment of the PCC and middle temporal gyrus (MTG) (Cha et al., 2013). In the PCC, MTG, and MFG regions, MMSE scores showed significant positive and negative associations with FC (Cha et al., 2013; Liao et al., 2018). Therefore, from the above studies, it was evident that most of the FC disruptions of the DMN occurred in the PCC and MTG (Bai et al., 2009; Zhou et al., 2008). Nevertheless, as the disease progresses, the FC disturbances spread to other brain regions (Damoiseaux, Prater, Miller, & Greicius, 2012). Since the PCC and other DMN hubs are affected in AD and MCI, the DMN may serve as an important biomarker for the classification of patients with AD and MCI. A recent review paper by Badhwar et al. that was published in 2017, studied various patterns of rs-fMRI dysfunctions among patients with AD, however they did not report on the accuracy of the test to distinguish the disease state (Badhwar et al., 2017). Therefore, our objective is to evaluate the diagnostic performance of rs-fMRI in identifying FC abnormalities of the DMN, which can help to distinguish between patients with AD or MCI from the healthy population.

## 2 Methodology

### 2.1 Study design

The systematic review method was adopted to achieve the current study based on the principles and methods provided by the York University’s Centre for Reviews and Dissemination guideline (CfRa, 2009). And the finding of the review were reported based on PRISMA (Prepared Reporting Items for Systematic Reviews and Meta-Analyses) (Moher, Liberati, Tetzlaff, & Altman, 2009).

### 2.2 Search strategy

A preliminary search was conducted to check for existing reviews in the Cochrane central register, Centre for Reviews and Dissemination such as Database of Abstract of Reviews of Effectiveness, National Health Service Economic Evaluation Database and the Health Technology Assessment Database, Turning Research Into Practice (TRIP) Database and for any on-going reviews similar to this study. This review protocol has been registered with the International Prospective Register of Systematic Reviews (PROSPERO) with the registration number CRD42020181655.

PubMed and Scopus databases were searched for articles using a combination of keyword terms such as Alzheimer’s Disease “OR” AD “AND” Resting state fMRI “AND” Default mode network “OR” DMN “AND” Sensitivity “AND” Specificity’ “OR” ‘Mild Cognitive Impairment “OR” MCI “AND” Resting state fMRI “AND” Default mode network “OR” DMN “AND” Sensitivity “AND” Specificity. We sourced for relevant published articles through 27, March 2020. The combined articles obtained from the search were screened for duplicates and the resultant articles underwent further screening as highlighted in the subsequent sections.

### 2.3 Criteria for study selection

#### 2.3.1 Inclusion criteria

The review paper included published original articles that met the following criteria: peer-reviewed articles written in the English language, the articles were sourced from journal publications until 27^th^, March 2020, and the articles included were observational studies of human subjects; which included case-control, cohort and cross-sectional studies utilizing rs-fMRI and the DMN to quantify and correlate FC between AD or MCI with healthy controls. Furthermore, the articles must have used established AD or MCI diagnostic criteria, e.g. Diagnostic and Statistics Manual of Mental Disorders IV or V (DSM-4 or DSM-5)(American Psychiatric Association, 2013) or the revised National Institute of Neurological and Communicative Disorders and Stroke and Alzheimer’s Disease and Related Disorders Association (NINCDS-ADRDA) (American Psychiatric Association, 2013).

#### 2.3.2 Exclusion criteria

We excluded articles by the following criteria: (a) Review articles (b) case reports (c) case series (d) articles written in foreign languages i.e. other than the English language (e) animal studies (f) articles with studies using imaging tools other than rs-fMRI, e.g. structural MRI, EEG, MEG or PET.

### 2.4 Data extraction

We conducted the literature search using the databases mentioned above. Two of the co-authors reviewed and independently screened the articles from the search results based on the titles and abstracts for potential inclusion into this review. Only the final screened articles agreed upon by both the authors were considered for the manuscript synthesis. In accordance with the PRISMA protocol, data extracted from each primary study included: author, year, country, number of subjects (patients and controls), age of the subjects, MMSE scores, rs-fMRI imaging protocol and analysis method, sensitivity scores, and specificity scores.

### 2.5 Quality Assessment

The quality of the methodology of the primary studies used in this review was assessed using Quality Assessment of Diagnostic Accuracy Study (QUADAS) tool (Beynon et al., 2012). The tool is designed with 14 questions to determine the risk of bias and applicability in terms of patient selection, index test, reference standard, and flow and timing domains. Based on the 13 questions, which served as a reference guide, each domain was rated low, high, or unclear (Appendix 1). The question “is the timing between reference standard and index test short enough to be reasonably sure that the target condition did not change between the two tests” cited from (Zhang et al., 2012), was removed because it does not fit the purpose of our review. A study was declared to have low risk of bias in a domain if many of the questions were positively scored for that domain (Zhang et al., 2012).

## 3 Results

### 3.1 Search results

The summary of the literature search results is in Figure 1. Out of the 225 primary articles obtained for AD studies, 14 duplicates were discarded. Then, 194 non-relevant articles were removed after screening based on titles and abstracts. Furthermore, eight articles were removed during the screening of full texts because of non-compliance with our study protocol. Out of the 165 primary articles related to MCI, we removed 7 duplicates and 147 non-relevant articles during screening. Finally, 8 articles were removed because they failed to conform to the study inclusion criteria. As a result, 12 articles met our inclusion criteria (9 AD and 3 MCI articles) and were included in the quantitative synthesis. In view of one study by Koch et al., 2012, having met the inclusion criteria for both AD and MCI, thus the final number of the included articles were 11 (Balthazar et al., 2014; Dai et al., 2012; de Vos et al., 2018; Koch et al., 2012; Krajcovicova et al., 2017; Li et al., 2012; X Miao et al., 2011; Park et al., 2017; Wang et al., 2013; Yokoi et al., 2018; Weimin Zheng et al.,2019).

**Figure 1:**
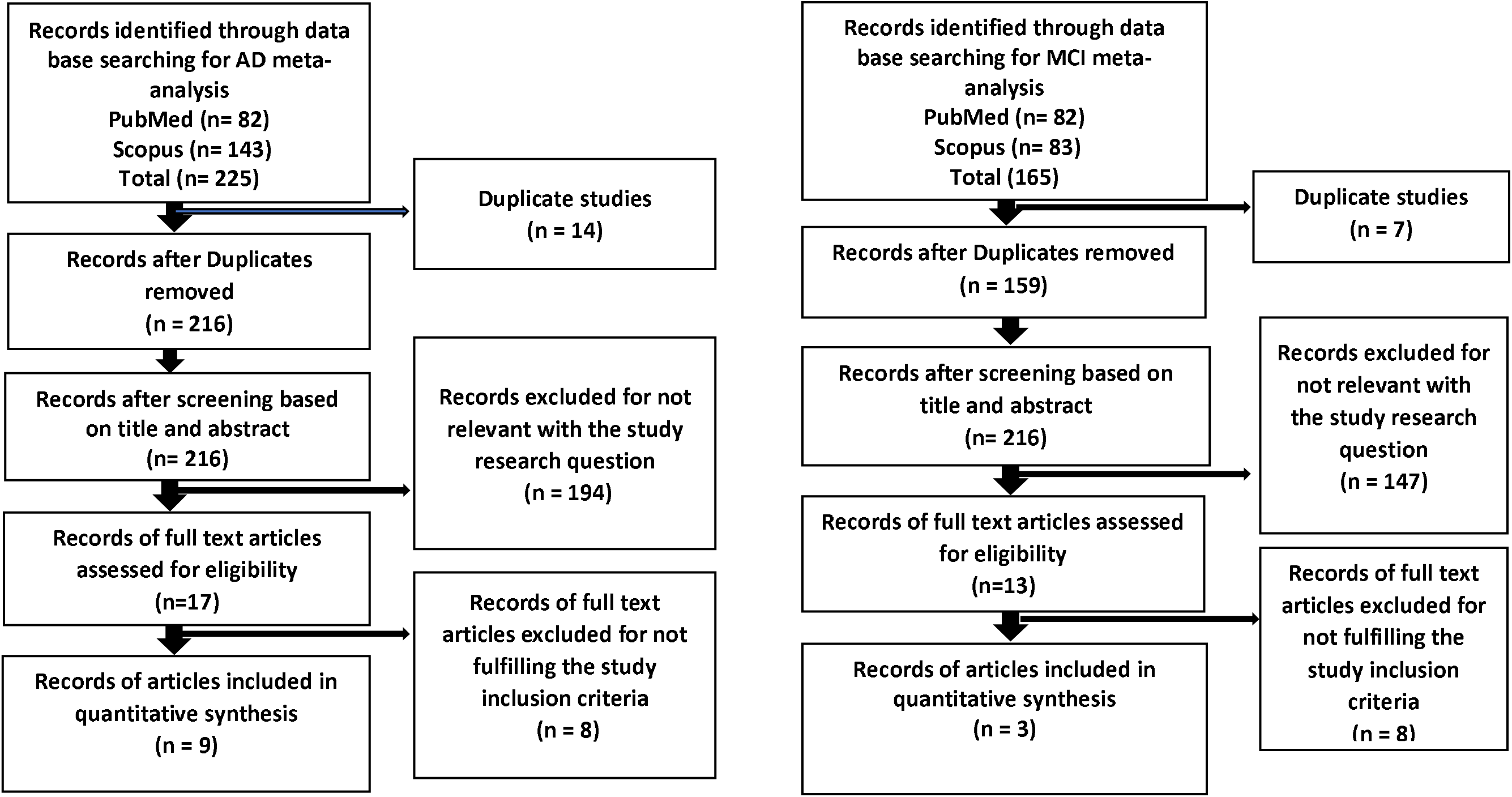
PRISMA flowchart summarizing the study selection process

### 3.2 Features of the primary studies included

Tables 1 and 2 summarize the main characteristics of the selected articles, which had assessed the rs-fMRI diagnostic performance for detecting DMN abnormalities among AD and MCI subjects. The majority (64%) of these articles had ≤ 20 subjects per group. Five articles (46%) used SBA and 3 studies (27%) used ICA type of analysis. While 1 article (9%) utilized both SBA and ICA methods, 2 articles (18%) used GTA as their method of analysis (Table 3). Interestingly, large percentage of these studies (55%) were conducted in Asia, with China having 5 out of the 7 studies from the region, the other two being from Japan and Korea, respectively. Three studies were performed in European countries such as Netherlands, Czech Republic, and Germany, respectively. Lastly, a study was carried out in Brazil too as shown in Table 1. As for the rs-fMRI parameters, most of the articles used 3-tesla scanners with a large percentage (64%) of the studies having repetition time (TR) of 2000 msec and echo time (TE) of 30 msec. Most of the studies had a scanning time of 10 minutes or less (Table 4).

**Table 1:** Demographic characteristics of the AD versus HC study participants.

| S/N | Author | Country | Subject |  |  |  |  |  |  |  |  |  |  |
| --- | --- | --- | --- | --- | --- | --- | --- | --- | --- | --- | --- | --- | --- |
|  |  |  | AD |  |  |  |  |  | HC |  |  |  |  |
|  |  |  | T | N | M | F | Age (SD) | MMSE (SD) | N | M | F | Age (SD) | MMSE (SD) |
| 1 | Balthazar et al. (2014) | Brazil | 48 | 22 | 6 | 16 | 73.40± 5.67 | 18.86± 4.68 | 26 | 6 | 20 | 71.03± 6.61 | 28.59± 1.86 |
| 2 | Dai, Z., et al. (2012) | China | 38 | 16 | 8 | 8 | 69.56 ± 7.65 | 18.50 ± 3.24 | 22 | 7 | 15 | 66.55 ± 7.67 | 28.59 ± 0.59 |
| 3 | de Vos, F., et al. (2018) | Netherlands | 250 | 77 | 31 | 46 | 68.6 ± 8.6 | 20.4 ± 4.5 | 173 | 74 | 99 | 66.1 ± 8.7 | 27.5 ± 1.8 |
| 4 | Koch et al., 2012 | Germany | 36 | 15 | 8 | 7 | 76.4± 10.3 | - | 21 | 10 | 11 | 68.6± 7.3 | - |
| 5 | Li, R., et al. (2012) | China | 21 | 15 | 6 | 9 | 64 ± 8.27 | 12 | 16 | 7 | 9 | 65 ± 9.20 | 29 |
| 6 | Park et al., (2017) | Korea | 63 | 41 | - | - | 71.2 ± 7.5 | 17.2 ± 5.4 | 22 | - | - | 67.9 ± 4.5 | 29.3 ± 1.6 |
| 7 | Zheng et al. (2019 | China | 70 | 40 | 18 | 22 | 65 ± 10 | 14.00 ± 6 | 30 | 15 | 15 | 64 ± 8 | 28.00 ± 2 |
| 8 | Miao, X., et al. (2011) | China | 31 | 15 | 6 | 8 | 64 ± 8.27 | 12 | 16 | 7 | 9 | 65 ± 9.20 | 29 |
| 9 | Yokoi, T., et al. (2018) | Japan | 47 | 23 | 4 | 19 | 68.6 ± 7.8 | 23.6 ± 2.8 | 24 | 8 | 16 | 65.4 ± 7.3 | 29.4 ± 1.0 |

T= total number of both AD and healthy control subjects, N= total number of subjects in either group M= male F= female, AD =Alzheimer's disease, HC = Healthy control

**Table 2:**
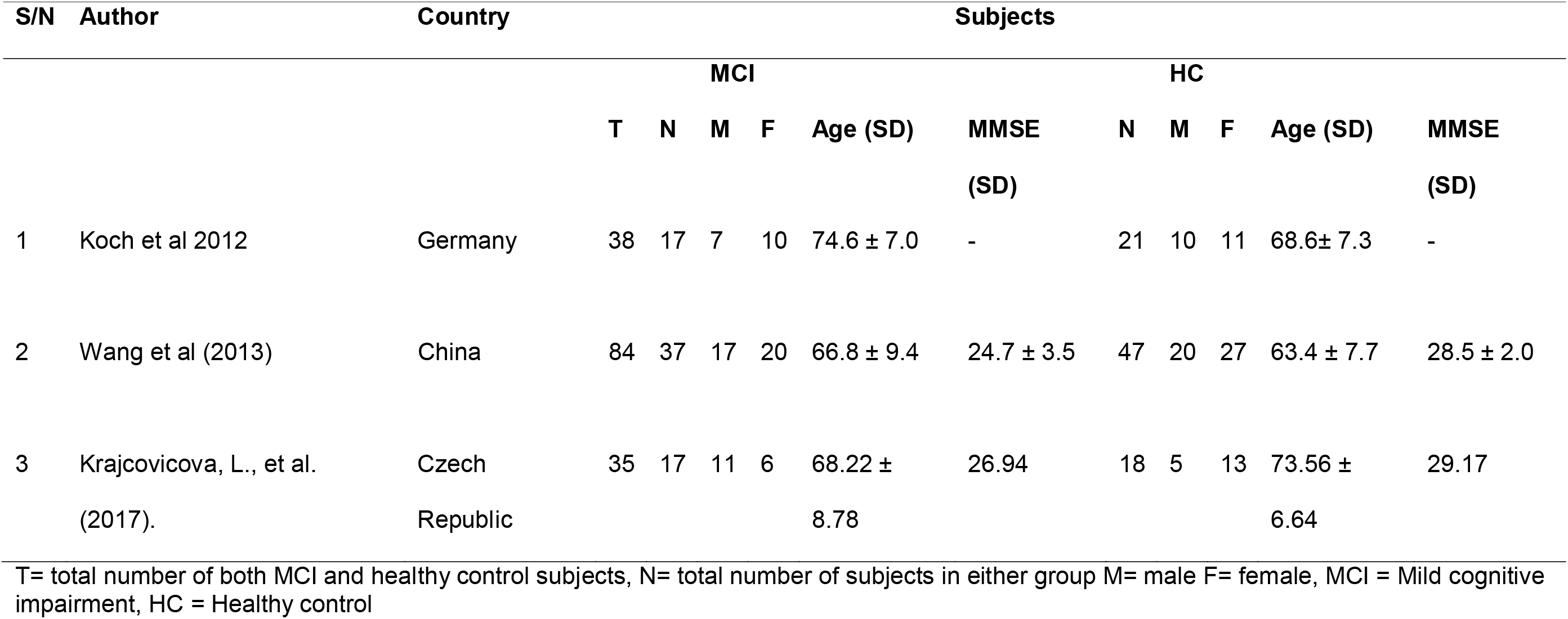
Sociodemographic of the MCI versus HC participants.

**Table 3:**
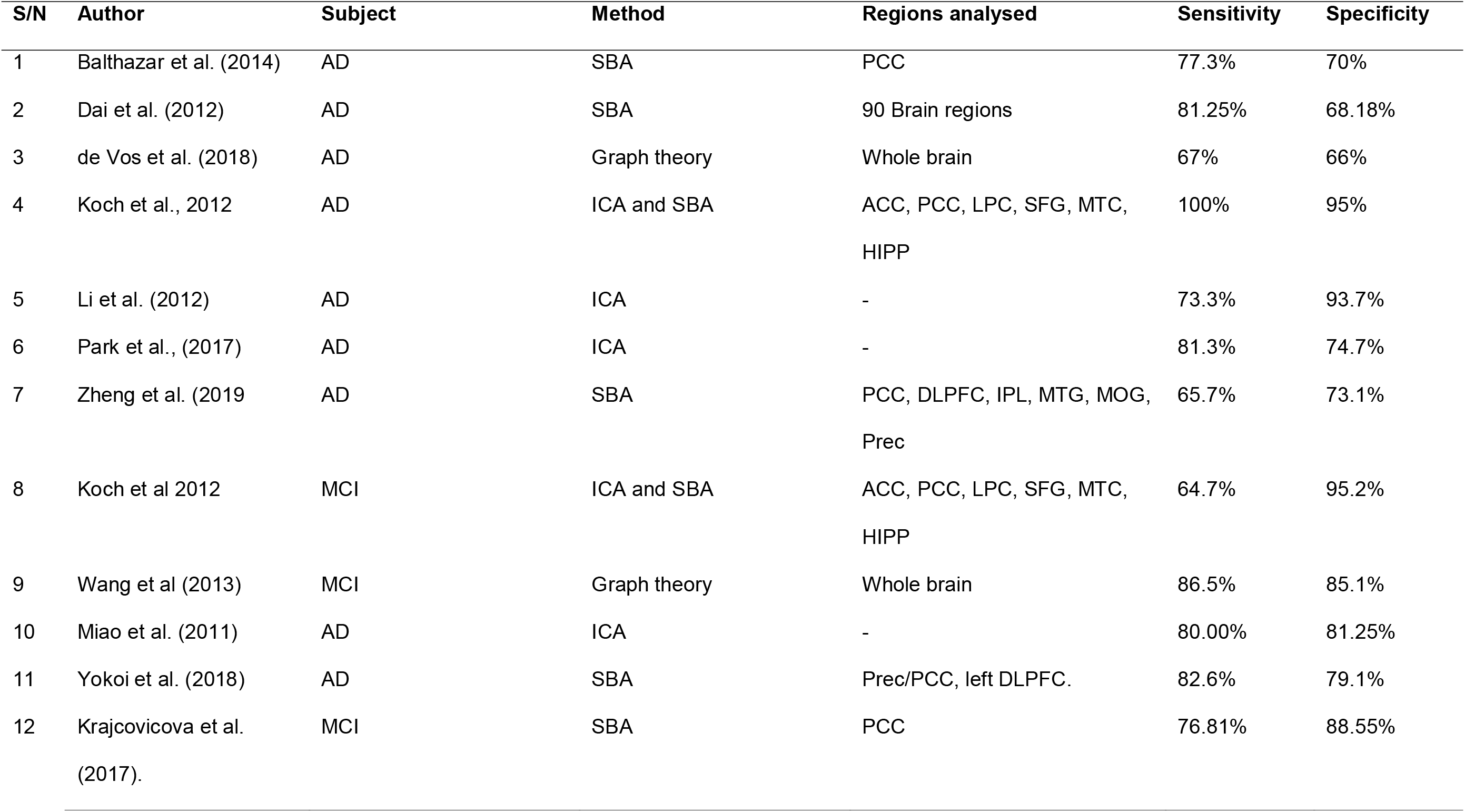
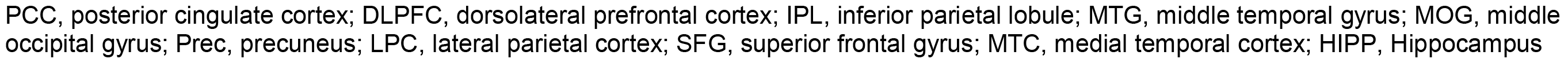
Regions of the DMN analysed using seed-based correlations.

**Table 4:** fMRI parameters of the selected studies.

| S/N | Author | Scanner<br>(T) | TR<br>(ms) | TE<br>(ms) | Other properties |
| --- | --- | --- | --- | --- | --- |
| 1 | Balthazar et al. (2014) | 3 P | 2000 | 30 | Field of view (FOV)= 240 × 240, axial slice= 40, volume= 300, Acquisition time (TA)= 10 mins, voxel size= 3×3×3 mm<sup>3</sup> |
| 2 | Dai, Z., et al. (2012) | 3 S | 2000 | 40 | Flip angle (FA)=90°; FOV=24 cm; matrix=64×64; slices=28; thickness=4 mm; voxel size=3.75×3.75×4 mm<sup>3</sup>; gap=1 mm; TA=478 s |
| 3 | de Vos, F., et al. (2018) | 3 S | 3000 | 30 | FA = 90°, axial slices= 40, voxel size of 3×3×3 mm, volume = 150 |
| 4 | Koch et al., 2012 | 3 S | 3000 | 30 | FA= 90°, voxel size = 3×3×4 mm<sup>3</sup>, imaging matrix= 64 × 64, FOV= 192 ×192 mm<sup>2</sup>, number of slices= 28, volumes=120, TA= 6 mins |
| 5 | Li, R., et al. (2012) | 3 S | 2000 | 30 | axial slices= 20 slice thickness= 6 mm, gap= 0 mm, FOV= 256 × 256 mm<sup>2</sup>; matrix size =64 × 64, FA= 85°; volume= 250 |
| 6 | Park et al. (2017) | 3 P | 3000 | 30 | FA= 90°; field of view= 212 mm; matrix= 64 x 64; slice thickness= 3 mm; gap= 0mm; number of slices= 47; volume= 140; TA= 7 minutes 3 seconds |
| 7 | Zheng et al. (2019) | 3 S | 2000 | 40 | FA= 90°, FOV = 24 cm, Image matrix = 64 × 64, slice number = 33, thickness = 3 mm, gap = 1 mm |
| 8 | Wang et al (2013) | 3 S | 2000 | 40 | FA= 90°; number of slices = 28; slice thickness = 4 mm; gap = 1 mm; voxel size = 4×4×4 |
|  |  |  |  |  | mm<sup>3</sup>; matrix = 64x64); Volume= 239; TA= 478 seconds |
| 9 | Miao, X., et al. (2011) | 3 S | 2000 | 30 | FA, 90°; voxel size = 3.13 × 3.13 × 3.6 mm<sup>3</sup>; FOV = 256 × 256 mm; TA = 300 |
| 10 | Yokoi, T., et al. (2018) | 3 S | 2500 | 30 | FA = 80°, number of slices = 39, 3-mm thickness, FOV= 192 mm, matrix = 64 × 64 mm, TA = 8 min |
| 11 | Krajcovicova, L., et al. (2017). | 1.5 S | 2050 | 50 | FA= 90°, FOV= 240 mm, matrix size = 64 × 64, slice thickness = 5 mm, volume = 250 |

P= Philips T= tesla S= Siemens

### 3.3 Quality of studies included

A summary of the assessment of risk of bias and applicability of the primary articles included in our review is depicted in Table 5. Most of the articles (73%), had a clear statement on patient selection procedures, hence were of low risk for attrition bias. All the included articles reported a clear statement on the index test, therefore had low risk of bias for this domain. In the reference standard domain, two articles (18%) ranked high risk for bias because they either did not report on how the reference standard was conducted or they did not use a reference standard. Finally, only one article had a high risk of bias in the flow and timing domain because additional subjects who did not follow the same screening procedures had significant age differences compared with the rest of the study subjects. For applicability, two articles (18%) in the patient selection and three articles (27%) in the reference standard were ranked as having a high risk of bias, either due to insufficient information or not using a reference standard. In general, most articles had overall low risk of bias and good applicability.

**Table 5:** Risk of bias Assessment.

| Study | Risk of bias |  |  |  | Applicability concerns |  |  |
| --- | --- | --- | --- | --- | --- | --- | --- |
|  | Patient | Index | Reference | Flow and | Patient | Index | Reference |
|  | Selection | test | standard | timing | Selection | test | standard |
| Balthazar et al., (2014) | L | L | L | L | H | L | L |
| Dai et al., (2012) | L | L | L | L | L | L | L |
| de Vos et al., (2018) | L | L | L | L | L | L | L |
| Koch et al., 2012 | H | L | L | H | L | L | H |
| Li et al. (2012) | H | L | H | L | L | L | H |
| Park et al., (2017) | L | L | L | L | L | L | L |
| Zheng et al. (2019) | L | L | L | L | L | L | L |
| Wang et al (2013) | L | L | L | L | L | L | L |
| Miao et al., (2011) | H | L | H | H | H | L | H |
| Yokoi et al., (2018) | L | L | L | L | L | L | L |
| Krajcovicova et al.,<br>(2017) | L | L | L | L | L | L | L |

Key H-high risk L-low risk U-unclear risk

### 3.4 Diagnostic Power of fMRI to identify DMN abnormalities in the Classification of AD

Nine articles included in our review demonstrated the diagnostic power of rs-fMRI in detecting DMN abnormalities for distinguishing AD patients from HCs, based on the brain regions that demonstrated weaker FC (Table 3 and 6) (Balthazar et al., 2014; Dai et al., 2012; de Vos et al., 2018; Koch et al., 2012; Li et al., 2012; Miao et al., 2011; Park et al., 2017; Yokoi et al., 2018; Zheng et al., 2019). Balthazar et al., (2014) evaluated the effect of FC of the DMN, namely impaired FC of the PCC together with cortical atrophy in discriminating 22 patients with mild AD, with 26 age and gender matched HC subjects. The authors found a moderate diagnostic power in the FC of the DMN (77.3% sensitivity and 70% specificity) between patients with mild AD and HCs. This indicates that even at the early stage of the AD, DMN moderately differentiate AD patients from HCs. Dai et al., (2012) performed discriminative analysis using FC, ReHo, ALFF and structural MRI grey matter density (GMD) to differentiate patient with mild AD from HCs. Some DMN nodes (including PCG, mPFC, hippocampus (HIPP) and parahippocampus) were found to be the most prominent distinguishing feature between patients with mild AD and HC. Additionally, in this study, the diagnostic power was 81.25% sensitivity and 68.18% specificity suggesting a high discriminating power of DMN FC among the patients with AD. De Vos et al., (2018) assessed the DMN impairments in AD patients, in a sample of 70 AD and 173 HCs, using GTA to analyse the FC. They reported a low diagnostic power of DMN between AD and controls with 66% and 67% sensitivity and specificity, respectively. Here, GTA analysis resulted into a low diagnostic power of DMN FC in differentiating AD patients from HCs.

Koch et al., (2012) reported 100% sensitivity and 95% specificity when they combined ICA and SBA to determine the diagnostic power of rs-fMRI in distinguishing DMN abnormalities among 15 AD patients and 21 HCs. This is an indication that a combination of two methods of analysis yielded a better diagnostic power of DMN between AD and HCs. Li et al., (2012) reported that the PCC can serve as a sensitive (73.3%) and specific (93.7%) biomarker for discriminating AD from HC, as shown in ICA analysis of 15 AD patients and 16 HCs. As indicated, a region of DMN (PCC) was able to discriminate diseased from HCs. A particular article in our review used Granger causality analysis to study the FC of the DMN among 15 AD and 16 HC subjects, in which they identified impaired connectivity in various DMN hubs among AD subjects (80% sensitivity and 81.25% specificity) (Xiaoyan Miao, Wu, Li, Chen, & Yao, 2011). Their result also demonstrated the high diagnostic power of DMN in distinguishing AD from HCs. Furthermore, in their attempt to evaluate the diagnostic power of MRI to detect cortical thickness and DMN FC for the classification of 41 AD patients and 22 HCs, Park et al., (2017) concluded that both parameters were significant biomarkers of AD, with the latter having 81.3% sensitivity and 74.7% specificity to classify AD from HCs.

In evaluating the difference between patients with early AD and HCs, Yokoi et al., (2018) compared the spatial retention of an amyloid marker, 18F-THK5351 and the DMN FC. It was noted that the Prec/ PCC is a specific hub for retention of 18F-THK5351, and that the disruption of both the PCC and the DLPFC account for fMRI diagnostic values of 82.6% sensitivity and 79.1% specificity. Zheng et al., (2019) reported the presence of disruption of the FC of the DMN and cerebral blood flow in the brain regions of 40 AD patients compared with 30 HC: namely in the PCC, DLPFC, inferior parietal lobule (IPL), MTG, MOG and Prec regions. It was observed that the disruption of the FC of the DMN discriminated AD from HCs, with a sensitivity of 65.7% and a specificity of 73.1%. This particular result showed lower discriminating power of fMRI in the classification of AD patients compared to the previously reported results. Furthermore, this drop in FC had a significant positive relationship with the decrease in the patients’ MMSE test scores (Zheng et al., 2019).

### 3.5 Diagnostic Power of fMRI in identifying DMN abnormalities in the Classification of MCI

Three articles that met our inclusion criteria for studying MCI subjects (Koch et al., 2012b; Krajcovicova et al., 2017; Wang et al., 2013), reported the diagnostic power of rs-fMRI in detecting impaired DMN FC. Koch et al., (2012) indicated that fMRI identified impaired FC of the DMN and correctly classified 17 MCI patients from 21 HCs. By using the combined methods of ICA and SBA, this yielded a sensitivity of 86.5% and a specificity of 85.1%. This result shows that rs-fMRI has a high diagnostic power in distinguishing MCI from HCs especially when using mutliple comparators. Krajcovicova et al., (2017), by utilising the SBA method, reported a moderate to high diagnostic power of rs-fMRI to correctly classify MCI patients (sensitivity and specificity of 76.81% and 88.55%, respectively). This article evaluated 17 MCI and 18 HCs, and identified DMN abnormalities predominantly in the PCCUsing whole-brain GTA method on 37 MCI and 47 HCs, Wang et al., (2013) demonstrated that disruptions in several nodes of the DMN can act as a biomarker for classifying MCI patients, giving a sensitivity of 86.5% and 85.1%, respectively. In all three studies, rs-fMRI demonstrated high diagnostic power in classifying abnormalities in the DMN of MCI patients compared to the HCs.

**Table 6:**
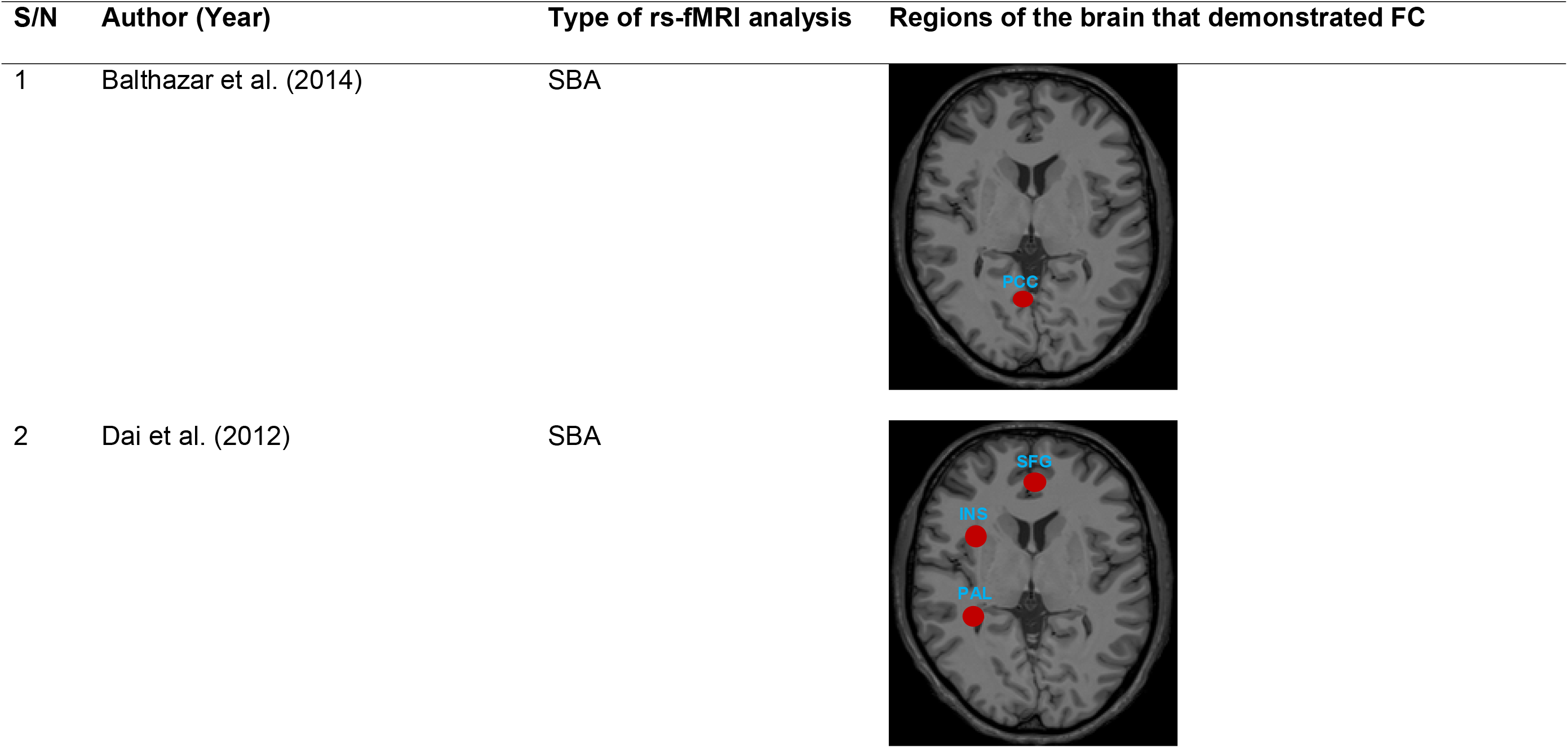
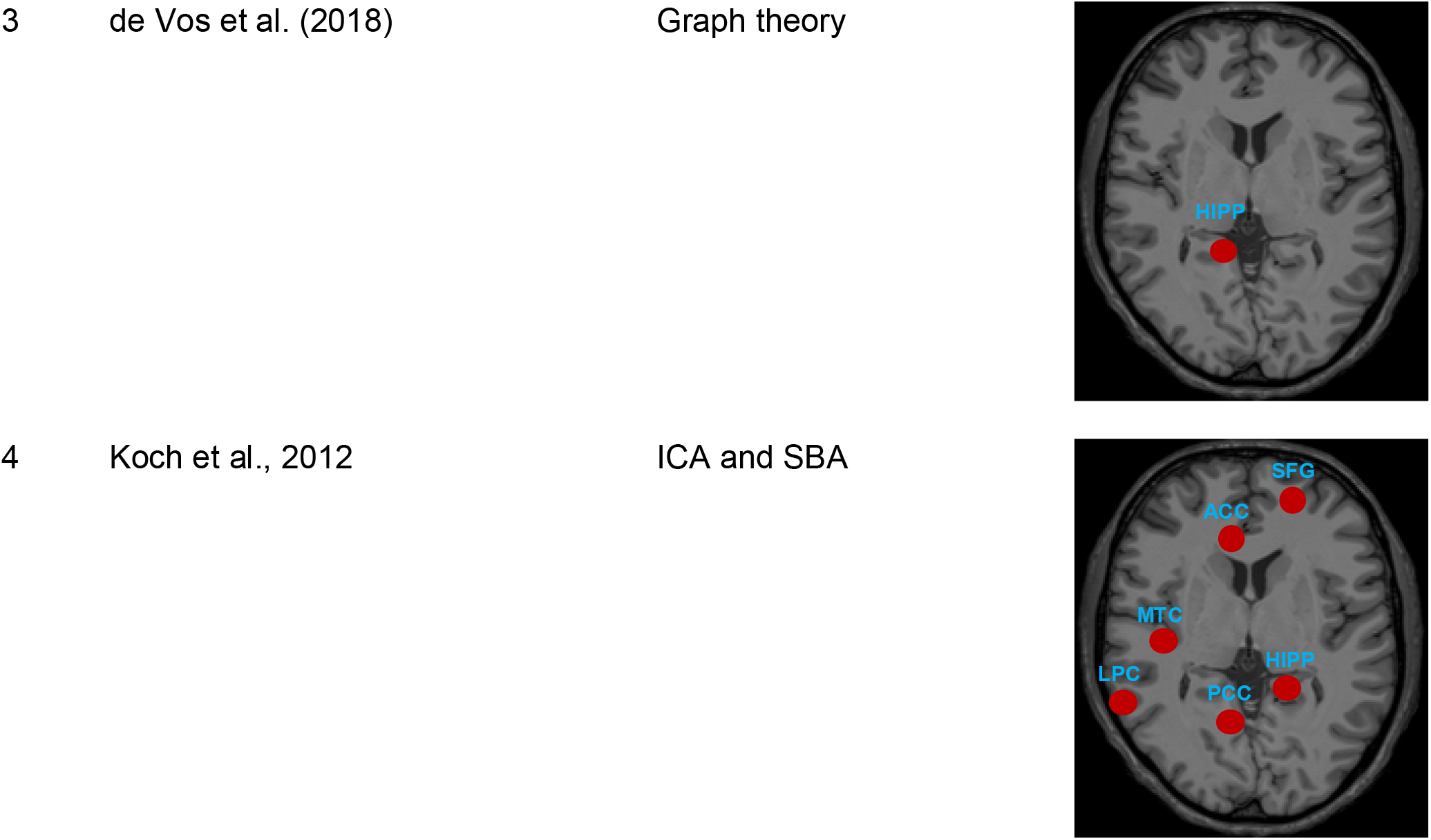
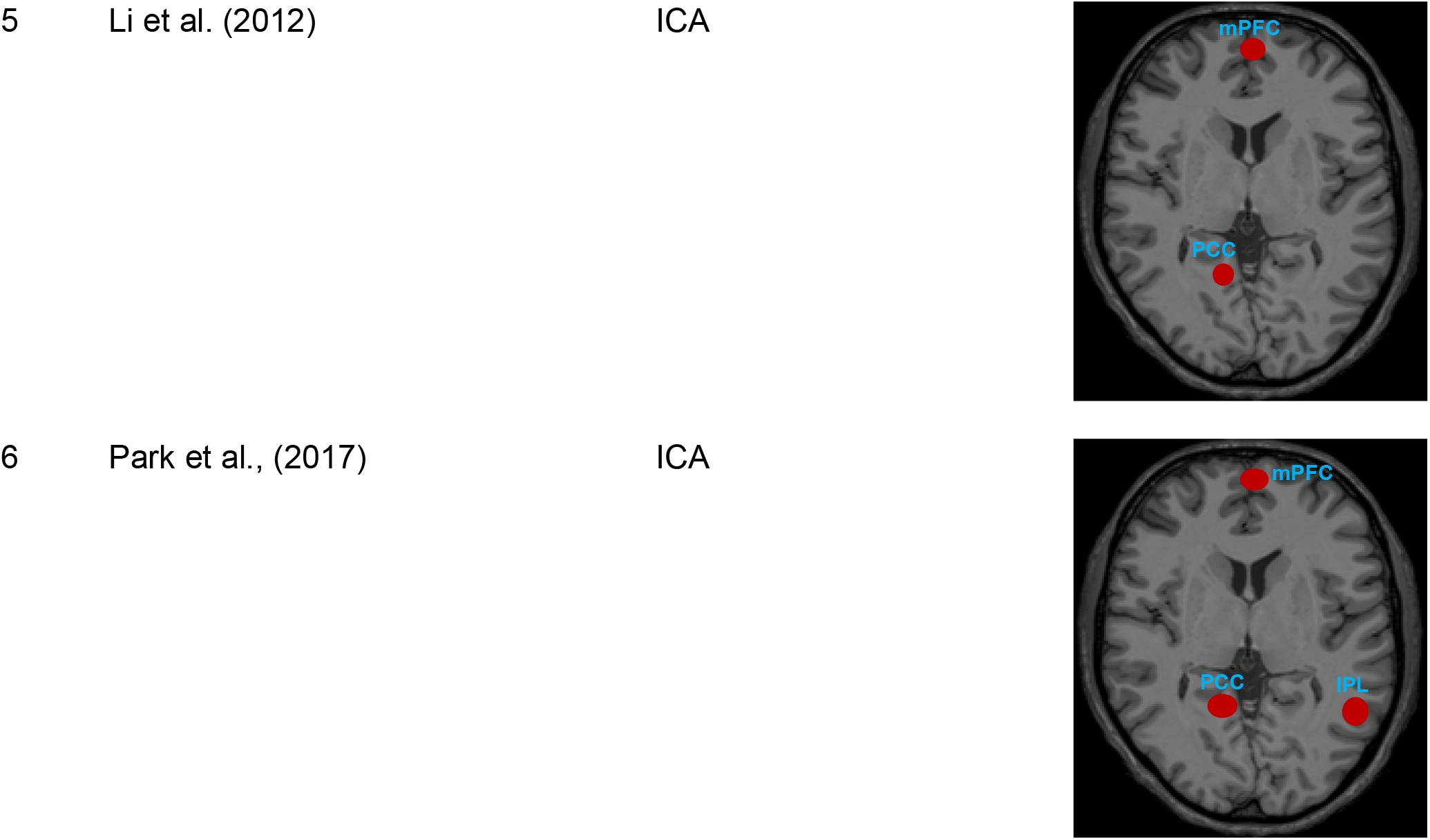
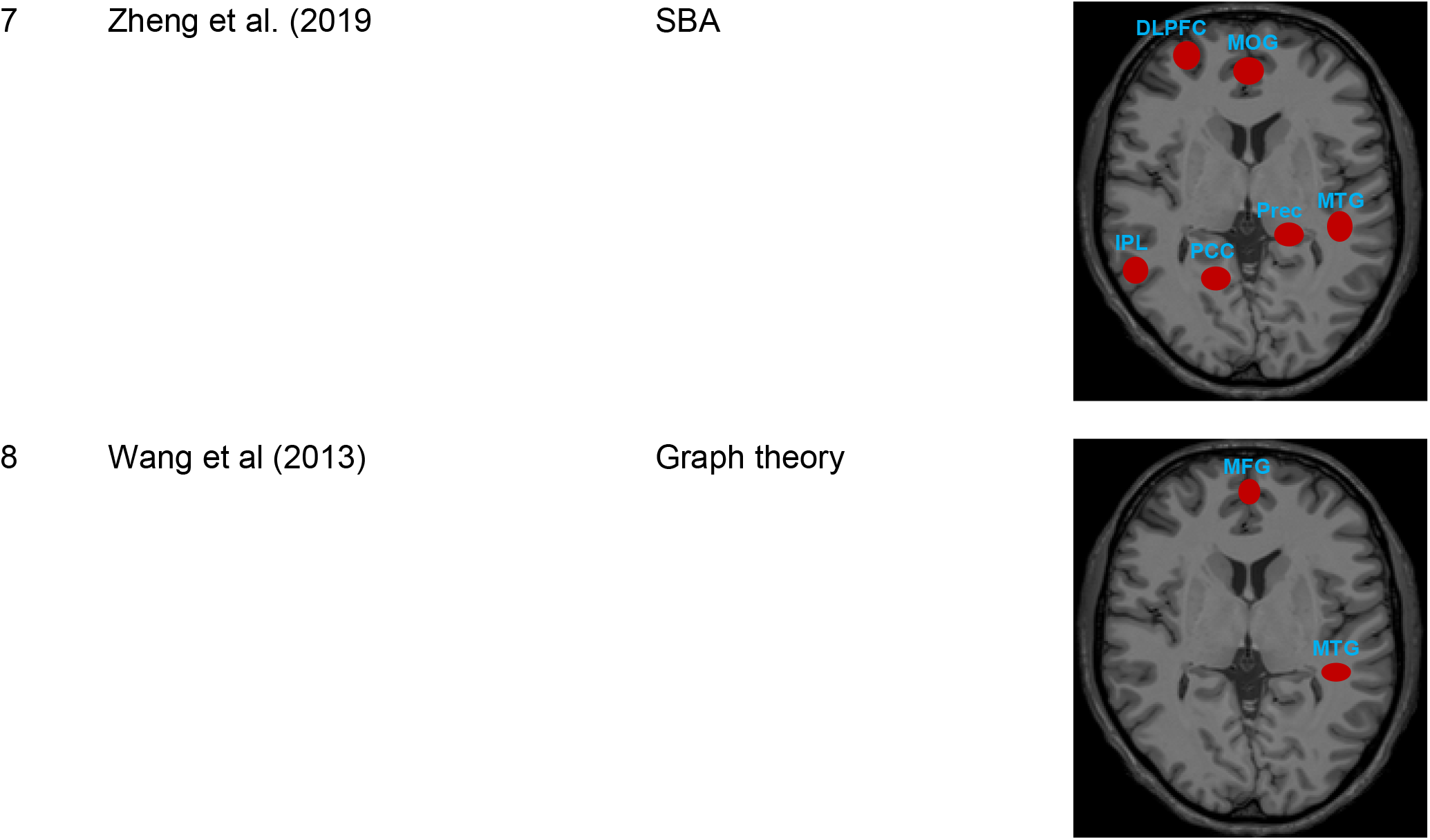
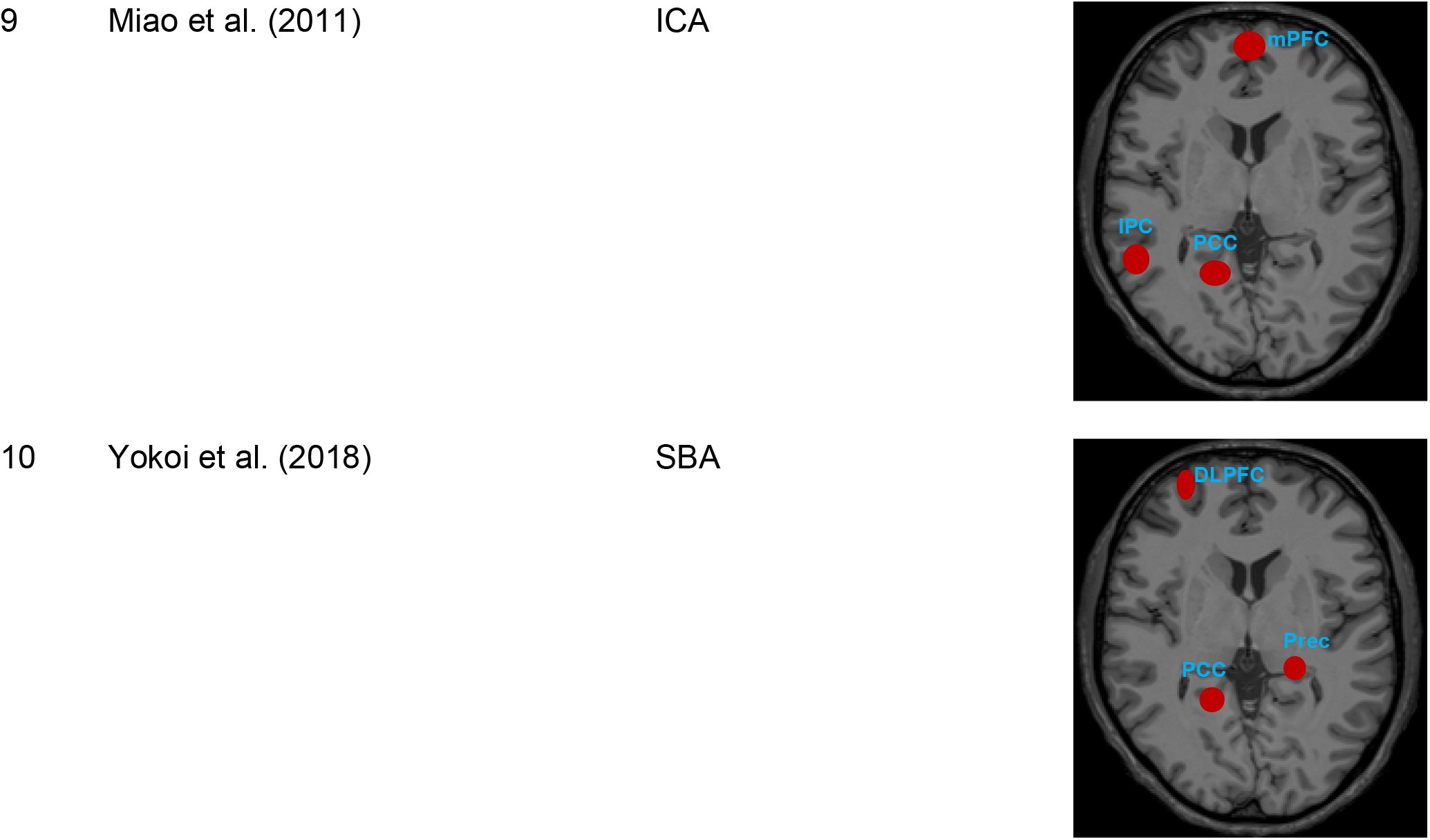
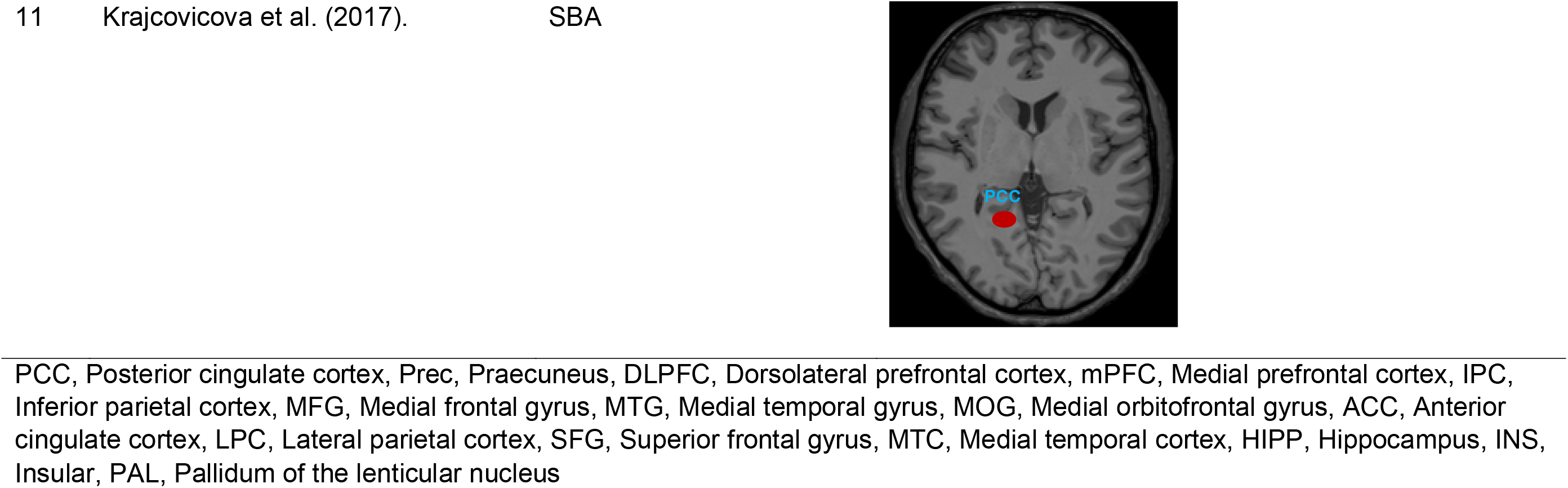
Regional FC on Bold fMRI in AD and MCI brain

## 4 Discussion

Although the FC of DMN has been explored as a biomarker for distinguishing patients with AD and MCI from HCs (Brier et al., 2012; Cha et al., 2013; Filippini et al., 2009; L Griffanti et al., 2015), no compiled review about its diagnostic power has been done prior to this. The impaired FC of DMN may be analysed using SBA, ICA, and GTA methods of analyses, and all these methods can be used to classify patients with AD and MCI.

To the best of our knowledge this is the first review to determine the diagnostic power of rs-fMRI to detect impairments in the FC of the DMN, for discriminating AD and MCI subjects from HCs. The articles included in this review reported variable diagnostic powers of rs-fMRI in characterizing AD and MCI patients, by using a variety of protocols, i.e. measurement of DMN FC alone (Koch et al., 2012b; H. Miao et al., 2016), DMN FC correlated with MRI-measured cortical thickness (Balthazar et al., 2014; Park et al., 2017), DMN FC measurements along with other resting-state measures such as DMN FC with PET/CT FC (Yokoi et al., 2018) and DMN FC with regional cerebral blood flow measurements (rCBF) (Zheng et al., 2019), respectively.

In differentiating AD patients from HCs, most of the primary articles used SBA analysis, all of which reported that AD patients had weaker FC between the PCC and other brain regions (Balthazar et al., 2014; Z Dai et al., 2019; Koch et al., 2012b; Yokoi et al., 2018; W Zheng et al., 2019). This biomarker, i.e. the PCC, gave an average sensitivity of 75.2% (ranging between 65.7% – 100%), and specificity of 74.9% (ranging between 70 – 95%) for distinguishing patients with AD, hence, indicating a moderate diagnostic power of DMN in differentiating AD patients from HCs. Even though SBA yielded good results, nevertheless, the applicability of solely evaluating the PCC is limited (Koch et al., 2012b) because the DMN has numerous hubs that are frequently disrupted in AD (Mohan et al., 2016; Wermke et al., 2008). Other regions of the DMN that were reported to have weaker FC in AD patients included the ACC, LPC, superior frontal gyrus (SFG), medial temporal cortex (MTC), HIPP, DLPFC, IPL, MTG, MOG and the Prec. The decreased FC among these regions are consistent with those reported in previous studies (Grieder et al., 2018; Griffanti et al., 2015; Rombouts et al., 2009). The deficient FC within regions of the DMN was correlated with MMSE scores, a global cognition and episodic memory measurement test (Balthazar et al., 2014; Yokoi et al., 2018). Moreover, this pattern of impaired DMN FC is in line with the course of early AD pathology, beginning from the MTG and involving the entorhinal cortex, HIPP, parahippocampus and fusiform gyri (Du et al., 2004; X. Li, Coyle, Maguire, Watson, & McGinnity, 2011). Advantageously, large-scale network (LSN) of rs-fMRI brain networks (DMN and dorsal attention network) can be studied using the ICA method. Articles that met our inclusion criteria, which utilized ICA (R. Li et al., 2012; X Miao et al., 2011; Park et al., 2017), reported that rs-fMRI had moderate to high diagnostic power to distinguished AD patients from HCs, having an average sensitivity of 78.2% (ranging between 73.3-81.3%) and an average specificity of 83.22% (ranging between 74.7-93.7%). The average sensitivity and specificity of these results were comparable with that of the SBA method. This is likely due to the same data being used for SBA was also used to construct independent component networks of the brain (Vemuri, Jones, & Jack, 2012). By combining both the ICA and SBA rs-fMRI methods and multivariate analysis to evaluate the ACC and PCC, Koch et al., 2012 achieved a high sensitivity and specificity, i.e.100% and 95%, respectively in discriminating AD from HC subjects. Apart from the SBA and ICA methods, GTA was an additional method employed to analyse fMRI findings in AD, which resulted in a moderate sensitivity of 67% and specificity of 66%, respectively (de Vos et al., 2018).

In evaluating patients with MCI, only one articles reported the PCC as the region of impaired DMN FC,, giving a sensitivity and specificity of 76.81% and 88.55%, respectively (Krajcovicova et al., 2017). In comparison, Koch et al., (2012), failed to produce any statistical difference between the FC of the DMN regions, i.e. in the ACC, PCC, LPC, SFG, MTC and HIPP, of the AD and HCs. They also reported a lower diagnostic power of rs-fMRI, i.e. sensitivity of 64.7% and specificity of 95.2%, respectively. Wang et al., (2013) used the GTA method to determine the diagnostic power of rs-fMRI to detected DMN FC abnormalities and to classify amnestic mild cognitive impairment (aMCI) from HCs. Their method achieved a high diagnostic power, with a sensitivity of 86.5% and specificity of 85.1%, respectively. Additionally, out of the three FC matrices utilised in their study, i.e. the global, nodal and FC strength, it was noted that the FC strength contributed the most to the power of the model for differentiating MCI from HCs.

In essence, rs-fMRI can detect impairment of the DMN FC and can serve to identify important anatomical biomarkers for discriminating AD and MCI patients from HCs. In particular, when combined with other parameters such as cortical thickness, rCBF, or analysed using combination of multivariate analysis, rs-fMRI has good diagnostic power for detecting AD and MCI.

### 4.1 Conclusion

The assessment of the DMN FC based on rs-fMRI analytic methods, has an excellent potential as a diagnostic tool for AD, particularly when using multivariate analysis to combine SBA and ICA methods of analyses. Nevertheless, the rs-fMRI protocols and analytical methods need to be more standardised to achieve uniformity in reporting improved diagnostic power.

### 4.2 Limitation and Recommendation

The relatively small sample size in the majority of the articles leads to a reduced power of the studies. Restrictions of the studies to only include subjects with early AD had to be made due to the constraints of performing the investigation on noncooperative patients with advanced AD. Furthermore, it is important to note that although MCI may occur as a prodromal condition to AD, it can also occur in vascular disease or even in cognitively healthy elderly persons without having progression to AD. Moreover, the conversion rate of MCI to AD is usually meagre. Therefore, only longitudinal studies can testify whether an MCI patient will develop to a full-blown AD, as opposed to identifying neural FC changes using a single timepoint rs-fMRI study. Additionally, there is need for improvement and standardization of rs-fMRI patient selection criteria, acquisition, image-processing and data analysis. The establishment of local population-based database of fMRI studies involving AD subjects can also help in improving the suitability of comparison.

## Data Availability

All data available are provided in the manuscript

## Acknowledgment

The research grant awarded to Associate Professor Dr Subapriya Suppiah by the Malaysian Ministry of Education, i.e. the Fundamental Research Grant Scheme (FRGS) with a grant number 5540244 and reference code number FRGS/1/2019/SKK03/UPM/02/4, helped support this research. (Geran Kementerian Pendidikan Malaysia, nombor kod rujukan FRGS/1/2019/SKK03/UPM/02/4).

## Conflict of Interest

The authors declare and report no conflict of interest.

## Author Contributions

SS conceptualised the study design. BI and NSN carried out the literature search, data extraction and quality assessment. BI wrote the manuscript first draft. SS, NI, MM, HAH and MIS edited the manuscript, verified the data, and provided critical feedback to help shape the research.

## Supplementary Table

**Supplementary Table:** list of the criteria used for assessing the methodological quality.

| S/N | Quality assessment | Positive response |
| --- | --- | --- |
| 1 | Was the spectrum of patients representative of the patients who will receive the test in practice? | Only AD and MCI patients were included and identified based on established diagnostic criteria either by DSM-IV or revised NINCDS–ADRDA |
| 2 | Were selection criteria clearly described? | It was clear how patients were selected for fMRI imaging |
| 3 | Is the reference standard likely to enable correct classification of the target condition? | Only clinical probable diagnosis of AD and MCI based on established diagnostic criteria either by DSM-IV or revised NINCDS–ADRDA is considered the reference standard |
| 4 | Did the whole sample or a random selection of the sample receive verification with a reference standard? | All patients received verification with DSM-IV or revised NINCDS–ADRDA is considered the reference standard |
| 5 | Did patients receive the same reference standard regardless of the index test result? | All patient received the same reference standard |
| 6 | Was the reference standard independent of the index test (the index test did not form part of the reference standard)? | fMRI did not form part of the reference standard |
| 7 | Was the execution of the index test described in sufficient detail to permit replication of the test? | The fMRI protocol including scanning time, type of scanner and acquisition mode were described in detail. |
| 8 | Was the execution of the reference standard described in sufficient detail to permit its replication? | Adequate information or citation on the reference standard was given |
| 9 | Were the index test results interpreted without knowledge of the results of the reference standard? | Interpretation of the fMRI results was done without the knowledge of the reference standard |
| 10 | Were the reference standard results interpreted without knowledge of the results of the index test? | Interpretation of the reference standard was done without the knowledge of the result of the index test |
| 11 | Were the same clinical data available when test results were interpreted as would be available when the test is used in practice? | All clinical and demographic characteristic of the patients was available when the test result was interpreted |
| 12 | Were uninterpretable and/or intermediate test results reported? | All the fMRI results (uninterpretable and/or intermediate) were reported |
| 13 | Were withdrawals from the study explained? | The reason for subject withdrawal was reported |

## Notes

### Competing Interest Statement

The authors have declared no competing interest.

### Author Declarations

It is a systematic review

## References

American Psychiatric Association. (2013). American Psychiatric Association, 2013. Diagnostic and statistical manual of mental disorders (5th ed.). In American Journal of Psychiatry. https://doi.org/10.1176/appi.books.9780890425596.744053

Badhwar, A., Tam, A., Dansereau, C., Orban, P., Hoffstaedter, F., & Bellec, P. (2017). Resting-state network dysfunction in Alzheimer’s disease: A systematic review and meta-analysis. Alzheimer’s and Dementia: Diagnosis, Assessment and Disease Monitoring, 8, 73–85. https://doi.org/10.1016/j.dadm.2017.03.007

Bai, F., Zhang, Z., Watson, D. R., Yu, H., Shi, Y., Yuan, Y., … Qian, Y. (2009). Abnormal Functional Connectivity of Hippocampus During Episodic Memory Retrieval Processing Network in Amnestic Mild Cognitive Impairment. Biological Psychiatry, 65(11), 951–958. https://doi.org/10.1016/j.biopsych.2008.10.017

Balthazar, M. L., de Campos, B. M., Franco, A. R., Damasceno, B. P., & Cendes, F. (2014). Whole cortical and default mode network mean functional connectivity as potential biomarkers for mild Alzheimer’s disease. Psychiatry Res, 221(1), 37–42. https://doi.org/10.1016/j.pscychresns.2013.10.010

Beynon, R., Sterne, J. A. C., Wilcock, G., Likeman, M., Harbord, R. M., Astin, M., … Whiting, P. (2012). Is MRI better than CT for detecting a vascular component to dementia? A systematic review and meta-analysis. BMC Neurology, 12, 33. https://doi.org/10.1186/1471-2377-12-33

Brier, M. R., Thomas, J. B., Snyder, A. Z., Benzinger, T. L., Zhang, D., Raichle, M. E., … Ances Dr., B. M. (2012). Loss of intranetwork and internetwork resting state functional connections with Alzheimer’s disease progression. Journal of Neuroscience. https://doi.org/10.1523/JNEUROSCI.5698-11.2012

Buckner, R. L., Sepulcre, J., Talukdar, T., Krienen, F. M., Liu, H., Hedden, T., … Johnson, K. A. (2009). Cortical hubs revealed by intrinsic functional connectivity: mapping, assessment of stability, and relation to Alzheimer’s disease. J Neurosci, 29(6), 1860–1873. https://doi.org/10.1523/jneurosci.5062-08.2009

CfRa, D. (2009). CRD’s guidance for undertaking reviews in health care. York Publishing Services Ltd, 32.

Cha, J., Jo, H. J., Kim, H. J., Seo, S. W., Kim, H. S., Yoon, U., … Lee, J. M. (2013). Functional alteration patterns of default mode networks: comparisons of normal aging, amnestic mild cognitive impairment and Alzheimer’s disease. Eur J Neurosci, 37(12), 1916–1924. https://doi.org/10.1111/ejn.12177

Chen, G., Ward, B. D., Xie, C., Li, W., Wu, Z., Jones, J. L., … Li, S. J. (2011). Classification of Alzheimer disease, mild cognitive impairment, and normal cognitive status with large-scale network analysis based on resting-state functional MR imaging. Radiology, 259(1), 213–221. https://doi.org/10.1148/radiol.10100734

Dai, Z, Lin, Q., Li, T., Wang, X., Yuan, H., Yu, X., … Wang, H. (2019). Disrupted structural and functional brain networks in Alzheimer’s disease. Neurobiol Aging, 75, 71–82. https://doi.org/10.1016/j.neurobiolaging.2018.11.005

Dai, Zhengjia, Yan, C., Wang, Z., Wang, J., Xia, M., Li, K., & He, Y. (2012). Discriminative analysis of early Alzheimer’s disease using multi-modal imaging and multi-level characterization with multi-classifier (M3). NeuroImage. https://doi.org/10.1016/j.neuroimage.2011.10.003

Damoiseaux, J. S., Prater, K. E., Miller, B. L., & Greicius, M. D. (2012). Functional connectivity tracks clinical deterioration in Alzheimer’s disease. Neurobiol Aging, 33(4), 828.e19-30. https://doi.org/10.1016/j.neurobiolaging.2011.06.024

De Luca, M., Smith, S., De Stefano, N., Federico, A., & Matthews, P. M. (2005). Blood oxygenation level dependent contrast resting state networks are relevant to functional activity in the neocortical sensorimotor system. Exp Brain Res, 167(4), 587–594. https://doi.org/10.1007/s00221-005-0059-1

de Vos, F., Koini, M., Schouten, T. M., Seiler, S., van der Grond, J., Lechner, A., … Rombouts, S. (2018). A comprehensive analysis of resting state fMRI measures to classify individual patients with Alzheimer’s disease. NeuroImage, 167, 6272. https://doi.org/10.1016/j.neuroimage.2017.11.025

Du, A. T., Schuff, N., Kramer, J. H., Ganzer, S., Zhu, X. P., Jagust, W. J., … Weiner, M. W. (2004). Higher atrophy rate of entorhinal cortex than hippocampus in AD. Neurology, 62(3), 422–427. https://doi.org/10.1212/01.wnl.0000106462.72282.90

Filippini, N., MacIntosh, B. J., Hough, M. G., Goodwin, G. M., Frisoni, G. B., Smith, S. M., … Mackay, C. E. (2009). Distinct patterns of brain activity in young carriers of the APOE-ε4 allele. Proceedings of the National Academy of Sciences of the United States of America. https://doi.org/10.1073/pnas.0811879106

Fox, M. D., & Raichle, M. E. (2007). Spontaneous fluctuations in brain activity observed with functional magnetic resonance imaging. Nat Rev Neurosci, 8(9), 700–711. https://doi.org/10.1038/nrn2201

Gili, T., Cercignani, M., Serra, L., Perri, R., Giove, F., Maraviglia, B., … Bozzali, M. (2011). Regional brain atrophy and functional disconnection across Alzheimer’s disease evolution. Journal of Neurology, Neurosurgery and Psychiatry. https://doi.org/10.1136/jnnp.2009.199935

Grieder, M., Wang, D. J. J., Dierks, T., Wahlund, L. O., & Jann, K. (2018). Default Mode Network Complexity and Cognitive Decline in Mild Alzheimer’s Disease. Front Neurosci, 12, 770. https://doi.org/10.3389/fnins.2018.00770

Griffanti, L, Dipasquale, O., Lagana, M. M., Nemni, R., Clerici, M., Smith, S. M., … Baglio, F. (2015). Effective artifact removal in resting state fMRI data improves detection of DMN functional connectivity alteration in Alzheimer’s disease. Front Hum Neurosci, 9, 449. https://doi.org/10.3389/fnhum.2015.00449

Griffanti, Ludovica, Dipasquale, O., Laganà, M. M., Nemni, R., Clerici, M., Smith, S. M., … Baglio, F. (2015). Effective artifact removal in resting state fMRI data improves detection of DMN functional connectivity alteration in Alzheimer’s disease. Frontiers in Human Neuroscience. https://doi.org/10.3389/fnhum.2015.00449

Jiang, T., He, Y., Zang, Y., & Weng, X. (2004). Modulation of functional connectivity during the resting state and the motor task. Hum Brain Mapp, 22(1), 63–71. https://doi.org/10.1002/hbm.20012

Koch, W., Teipel, S., Mueller, S., Benninghoff, J., Wagner, M., Bokde, A. L., … Meindl, T. (2012a). Diagnostic power of default mode network resting state fMRI in the detection of Alzheimer’s disease. Neurobiol Aging, 33(3), 466–478. https://doi.org/10.1016/j.neurobiolaging.2010.04.013

Koch, W., Teipel, S., Mueller, S., Benninghoff, J., Wagner, M., Bokde, A. L. W., … Meindl, T. (2012b). Diagnostic power of default mode network resting state fMRI in the detection of Alzheimer’s disease. Neurobiology of Aging, 33(3), 466–478. https://doi.org/10.1016/j.neurobiolaging.2010.04.013

Krajcovicova, L., Barton, M., Elfmarkova-Nemcova, N., Mikl, M., Marecek, R., & Rektorova, I. (2017). Changes in connectivity of the posterior default network node during visual processing in mild cognitive impairment: staged decline between normal aging and Alzheimer’s disease. J Neural Transm (Vienna), 124(12), 1607-1619. https://doi.org/10.1007/s00702-017-1789-5

Lee, L., Harrison, L. M., & Mechelli, A. (2003). A report of the functional connectivity workshop, Dusseldorf 2002. NeuroImage. https://doi.org/10.1016/S1053-8119(03)00062-4

Li, R., Wu, X., Fleisher, A. S., Reiman, E. M., Chen, K., & Yao, L. (2012). Attention-related networks in Alzheimer’s disease: a resting functional MRI study. Hum Brain Mapp, 33(5), 1076–1088. https://doi.org/10.1002/hbm.21269

Li, X., Coyle, D., Maguire, L., Watson, D. R., & McGinnity, T. M. (2011). Gray matter concentration and effective connectivity changes in Alzheimer’s disease: A longitudinal structural MRI study. Neuroradiology. https://doi.org/10.1007/s00234-010-0795-1

Liao, Z. L., Tan, Y. F., Qiu, Y. J., Zhu, J. P., Chen, Y., Lin, S. S., … Yu, E. Y. (2018). Interhemispheric functional connectivity for Alzheimer’s disease and amnestic mild cognitive impairment based on the triple network model. Journal of Zhejiang University: Science B, 19(12), 924–934. https://doi.org/10.1631/jzus.B1800381

Liu, J., Zhang, X., Yu, C., Duan, Y., Zhuo, J., Cui, Y., … Liu, Y. (2016). Impaired Parahippocampus Connectivity in Mild Cognitive Impairment and Alzheimer’s Disease. J Alzheimers Dis, 49(4), 1051–1064. https://doi.org/10.3233/jad-150727

Miao, H., Chen, X., Yan, Y., He, X., Hu, S., Kong, J., … Qiu, B. (2016). Functional connectivity change of brain default mode network in breast cancer patients after chemotherapy. Neuroradiology, 58(9), 921–928. https://doi.org/10.1007/s00234-016-1708-8

Miao, X, Wu, X., Li, R., Chen, K., & Yao, L. (2011). Altered connectivity pattern of hubs in default-mode network with alzheimer’s disease: An granger causality modeling approach. PLoS ONE, 6(10). https://doi.org/10.1371/journal.pone.0025546

Miao, Xiaoyan, Wu, X., Li, R., Chen, K., & Yao, L. (2011). Altered connectivity pattern of hubs in default-mode network with alzheimer’s disease: An granger causality modeling approach. PLoS ONE. https://doi.org/10.1371/journal.pone.0025546

Mier, W., & Mier, D. (2015). Advantages in functional imaging of the brain. Frontiers in Human Neuroscience. https://doi.org/10.3389/fnhum.2015.00249

Mohan, A., Roberto, A. J., Mohan, A., Lorenzo, A., Jones, K., Carney, M. J., … Lapidus, K. A. B. (2016). The significance of the Default Mode Network (DMN) in neurological and neuropsychiatric disorders: A review. Yale Journal of Biology and Medicine.

Moher, D., Liberati, A., Tetzlaff, J., & Altman, D. G. (2009). Preferred reporting items for systematic reviews and meta-analyses: the PRISMA statement. PLoS Med, 6(7), e1000097. https://doi.org/10.1371/journal.pmed.1000097

Ouchi, Y., & Kikuchi, M. (2012). A review of the default mode network in aging and dementia based on molecular imaging. Reviews in the Neurosciences. https://doi.org/10.1515/revneuro-2012-0029

Park, J. E., Park, B., Kim, S. J., Kim, H. S., Choi, C. G., Jung, S. C., … Shim, W. H. (2017). Improved diagnostic accuracy of alzheimer’s disease by combining regional cortical thickness and default mode network functional connectivity: Validated in the alzheimer’s disease neuroimaging initiative set. Korean Journal of Radiology, 18(6), 983–991. https://doi.org/10.3348/kjr.2017.18.6.983

Rombouts, S. A., Damoiseaux, J. S., Goekoop, R., Barkhof, F., Scheltens, P., Smith, S. M., & Beckmann, C. F. (2009). Model-free group analysis shows altered BOLD FMRI networks in dementia. Hum Brain Mapp, 30(1), 256–266. https://doi.org/10.1002/hbm.20505

Rubinov, M., & Sporns, O. (2010). Complex network measures of brain connectivity: uses and interpretations. Neuroimage, 52(3), 1059–1069. https://doi.org/10.1016/j.neuroimage.2009.10.003

Vemuri, P., Jones, D. T., & Jack, C. R. (2012). Resting state functional MRI in Alzheimer’s disease. Alzheimer’s Research and Therapy. https://doi.org/10.1186/alzrt100

Wang, Y., Chen, K., Yao, L., Jin, Z., & Guo, X. (2013). Structural interactions within the default mode network identified by Bayesian network analysis in Alzheimer’s disease. PLoS ONE, 8(8), e74070. https://doi.org/10.1371/journal.pone.0074070

Wee, C. Y., Yap, P. T., Zhang, D., Denny, K., Browndyke, J. N., Potter, G. G., … Shen, D. (2012). Identification of MCI individuals using structural and functional connectivity networks. NeuroImage, 59(3), 2045–2056. https://doi.org/10.1016/j.neuroimage.2011.10.015

Wermke, M., Sorg, C., Wohlschläger, A. M., & Drzezga, A. (2008). A new integrative model of cerebral activation, deactivation and default mode function in Alzheimer’s disease. European Journal of Nuclear Medicine and Molecular Imaging. https://doi.org/10.1007/s00259-007-0698-5

Yokoi, T., Watanabe, H., Yamaguchi, H., Bagarinao, E., Masuda, M., Imai, K., … Sobue, G. (2018). Involvement of the precuneus/posterior cingulate cortex is significant for the development of Alzheimer’s disease: A PET (THK5351, PiB) and resting fMRI study. Frontiers in Aging Neuroscience, 10(OCT). https://doi.org/10.3389/fnagi.2018.00304

Zhang, S., Han, D., Tan, X., Feng, J., Guo, Y., & Ding, Y. (2012). Diagnostic accuracy of 18F-FDG and 11C-PIB-PET for prediction of short-term conversion to Alzheimer’s disease in subjects with mild cognitive impairment. International Journal of Clinical Practice. https://doi.org/10.1111/j.1742-1241.2011.02845.x

Zheng, D., Xia, W., Yi, Z. Q., Zhao, P. W., Zhong, J. G., Shi, H. C., … Pan, P. L. (2018). Correction to: Alterations of brain local functional connectivity in amnestic mild cognitive impairment. Transl Neurodegener, 7, 33. https://doi.org/10.1186/s40035-018-0140-x

Zheng, W, Cui, B., Han, Y., Song, H., Li, K., He, Y., & Wang, Z. (2019). Disrupted regional cerebral blood flow, functional activity and connectivity in Alzheimer’s disease: A combined ASL perfusion and resting state fMRI study. Frontiers in Neuroscience, 13(JUL). https://doi.org/10.3389/fnins.2019.00738

Zheng, Weimin, Cui, B., Han, Y., Song, H., Li, K., He, Y., & Wang, Z. (2019). Disrupted regional cerebral blood flow, functional activity and connectivity in Alzheimer’s disease: A combined ASL perfusion and resting state fMRI study. Frontiers in Neuroscience. https://doi.org/10.3389/fnins.2019.00738

Zhou, Y., Dougherty Jr., J. H., Hubner, K. F., Bai, B., Cannon, R. L., & Hutson, R. K. (2008). Abnormal connectivity in the posterior cingulate and hippocampus in early Alzheimer’s disease and mild cognitive impairment. Alzheimers Dement, 4(4), 265–270. https://doi.org/10.1016/jjalz.2008.04.006

